# Frontoparietal structural network disconnections correlate with outcome after a severe stroke

**DOI:** 10.1101/2024.03.22.24304660

**Authors:** Lukas Frontzkowski, Felix Fehring, Benedikt M. Frey, Paweł P. Wróbel, Antonia Reibelt, Focko Higgen, Silke Wolf, Winifried Backhaus, Hanna Braaß, Philipp J. Koch, Chi-un Choe, Marlene Bönstrup, Bastian Cheng, Götz Thomalla, Christian Gerloff, Fanny Quandt, Robert Schulz

## Abstract

Structural disconnectome analyses have provided valuable insights into how a stroke lesion results in widespread network disturbances and how these relate to deficits, recovery patterns, and outcome. Previous analyses have primarily focused on patients with relatively mild to moderate deficits. However, outcomes vary among survivors of severe strokes, and the mechanisms of recovery remain poorly understood. This study assesses the association between lesion-induced network disconnection and outcome after severe stroke. Thirty-eight ischaemic stroke patients underwent MRI brain imaging early after stroke and longitudinal clinical follow-up. Lesion information was integrated with normative connectome data to infer individual disconnectome profiles on a localized regional and region-to-region pathway level. Ordinal logistic regressions were computed to link disconnectome information to the modified Rankin Scale after 3-6 months. Disconnections of ipsilesional frontal, parietal and temporal cortical brain areas were significantly associated with a worse motor outcome after a severe stroke, adjusted for the initial deficit, lesion volume, and age. The analysis of the underlying pathways mediating this association revealed location specific results: For frontal, prefrontal and temporal brain areas, the association was primarily driven by relatively sparse intrahemispheric disconnections. In contrast, the ipsilesional primary motor cortex, the dorsal premotor cortex, and various parietal brain regions showed a remarkable involvement of either frontoparietal intrahemispheric or additionally interhemispheric disconnections. These results indicate that localized disconnection of multiple regions embedded in the structural frontoparietal network correlates with worse outcome after severe stroke. Specifically, primary motor and parietal cortices might gain a particular importance as they structurally link frontoparietal networks of both hemispheres. These data shed novel light on the significance of distinct brain networks for recovery after severe stroke.

## Introduction

Acute stroke treatment with thrombolysis and mechanical thrombectomy has significantly enhanced survival rates and overall outcome after ischemic stroke. However, particularly in patients exhibiting severe initial deficits, recovery is incomplete, and survivors are left with functional disabilities that critically impede private and professional independence ^1-3^. In recent years, structural brain imaging has offered invaluable insights into the recovery processes following stroke, aiming to explain the inter-subject variability in stroke patient outcomes ^4^. It not only sought to improve outcome modelling, ultimately to guide clinical decision making and rehabilitation strategies ^5^, but structural brain imaging also aimed to enhance our mechanistic understanding of how the brain and its networks respond to stroke lesions and to identify which structural properties may contribute to a favourable outcome ^6-8^. While investigations of specific tracts such as the corticospinal tract have dominated the field for years ^4^, a network perspective, including the analysis of lesion-induced disturbances of network connections, has been more recently incorporated with varying methodological approaches ^9,10^. Notably, the integration of simple lesion information into fine-grained normative healthy connectome data for indirect disconnectome-symptom mapping ^10^ may facilitate the transfer from neuroscience to potential clinical applications.

Patterns of network disconnectivity have been associated with post-stroke deficits in subacute stroke patients ^11^, with variability in recovery trajectories over time ^7,12-15^, with treatment gains during motor rehabilitation paradigms ^16^, and fatigue ^17^ or cognitive functions ^18^ in the chronic stage of recovery. Here, profiles of single regions disconnected from larger brain networks, and profiles of disturbed interconnecting pathways at a pairwise region-to-region level have been found to carry relevant information ^12^. Treatment-related insights have been provided by assessing effects of thrombolysis onto network disconnections ^19^ and topology ^20^, or the influence of tract-specific disconnections before thrombectomy for outcome inference ^21^. As one important limitation however, at least some of these disconnectome analyses have primarily focused on patients with rather mild to moderate deficits with initial National Institutes of Health Stroke Scale (NIHSS) scores ranging between 1 and 3 ^7,11,13,18^ or up to 7 ^12,20^. Data regarding severely impaired patients are limited. Though, particularly survivors of severe stroke are characterized by great variability in recovery patterns and uncertainty regarding underlying, supportive mechanisms on network neuroscience levels.

Therefore, the present work aimed to assess the relationship between lesion-induced network disconnection profiles and subsequent outcome after severe stroke. First-ever supratentorial ischemic stroke patients with severe deficits underwent clinical testing and brain imaging within two weeks after the event. Lesion information was integrated with normative connectome data to infer individual disconnectome profiles on a localized regional and region-to-region level. Ordinal logistic regression modelling was conducted to link disconnectome information to the global disability as the clinical outcome, operationalized by the modified Rankin Scale (mRS) after 3-6 months. We hypothesized that regional disconnection particularly of ipsilesional frontal and parietal brain regions ^22^ from larger networks will be significantly correlated with outcome variability. Frontal and parietal areas will differ in the extent to which specific intra- or interhemispheric disconnectivity will drive the overall region’s importance. Finally, when compared to previous publications studying less impaired cohorts, the present findings might allow novel insights into severe stroke recovery ^3,23^, not only aiming to improve current prediction models, but also paving the way towards patient-tailored innovative treatment strategies including non-invasive brain stimulation ^24^.

## Methods

### Demographic data

The datasets of 38 severely affected stroke patients from two independent, previously published observational studies (Cohort 1, C1 ^25^, Cohort 2, C2 ^26^) were included in this analysis. Acute stroke patients admitted to the University Medical Center Hamburg-Eppendorf were recruited according to following criteria: first-ever unilateral ischemic supratentorial stroke, upper extremity motor deficit involving hand function, no history of previous neurological or psychiatric illness and age ≥ 18 years. Magnetic resonance imaging (MRI) and clinical testing was performed in the first 3-14 days after stroke onset (time point T_1_). In the late subacute stage of stroke recovery (3 months post-stroke), clinical testing was repeated and defined as follow-up time point T_2_. A subset of patients was not eligible for clinical testing at timepoint T_2_ and thus, data was only available after 6 months. This approach has been repeatedly used in our previous analyses which also give precise information regarding the cohorts’ integration ^26-28^. In this analysis, initial symptom burden was operationalized by the NIHSS score at T_1_, global disability at T_2_ was quantified by mRS.

### Image acquisition

A 3T Skyra MRI scanner (Siemens Healthineers, Erlangen, Germany) equipped with a 32-channel head coil was used to obtain structural high-resolution T1-weighted images applying a 3-dimensional magnetization-prepared rapid gradient echo sequence (MPRAGE) [repetition time (TR) = 2500 ms, echo time (TE) = 2.12 ms, flip angle 9°, 256 coronal slices with a voxel size of 0.8 × 0.8 × 0.9 mm^3^, field of view (FOV) = 240 mm]. T2-weighted images were also acquired by using a fluid-attenuated inversion recovery sequence (TR = 9000 ms, TE = 86 ms, TI = 2500 ms, flip angle 150°, 43 transversal slices with a voxel size of 0.7 × 0.7 × 3.0 mm^3^, FOV = 230 mm) for stroke lesion delineation.

### Quantification of structural disconnection

Stroke lesions were semi-automatically delineated with SNAP-ITK ^29^ and available from our previous analyses ^26-28,30^. The resulting lesion masks were registered to a 1-mm^3^ Montreal Neurological Institute (MNI) template. To compute structural disconnection profiles for each subject, masks were analysed using the network modification NEMO2-toolbox which computes the disruption of white matter connections by overlaying an MNI-registered lesion mask onto a normative structural connectome derived from 420 healthy Human Connectome Project participants ^31^. In brief, a change of connectivity (ChaCo) measure is computed which resembles the extent of disrupted streamlines divided by total streamlines for either each voxel or an atlas-based parcellation. For our analyses, we applied the Brainnetome atlas as it is based on a connectional mapping approach and thus applicable for testing the extent of disconnection between brain regions ^32^. ChaCo values were computed on a regional level and a pairwise region-to-region level (i.e., addressing interconnecting pathways contributing to the connectivity of each region). In line with previous recommendations, all values below 0.02 were disregarded due to noise artefacts ^31^. To increase comparability between stroke patients, ChaCo values were flipped to the left side when the stroke site was in the right hemisphere. We excluded all regions with a critically skewed data distribution (i.e., skew either >1.3 or <-1.3). Out of 246 Brainnetome regions in total, 100 regions were eligible for further statistical analyses (Figure 1, Suppl. Figure 1 and Suppl. Table 1). For sensitivity analyses, two alternative skew thresholds (+/-1.0 and +/-1.5, respectively) were explored. However, the extent and distribution of eligible regions for modelling remained stable (Suppl. Figure 2).

**Figure 1:**
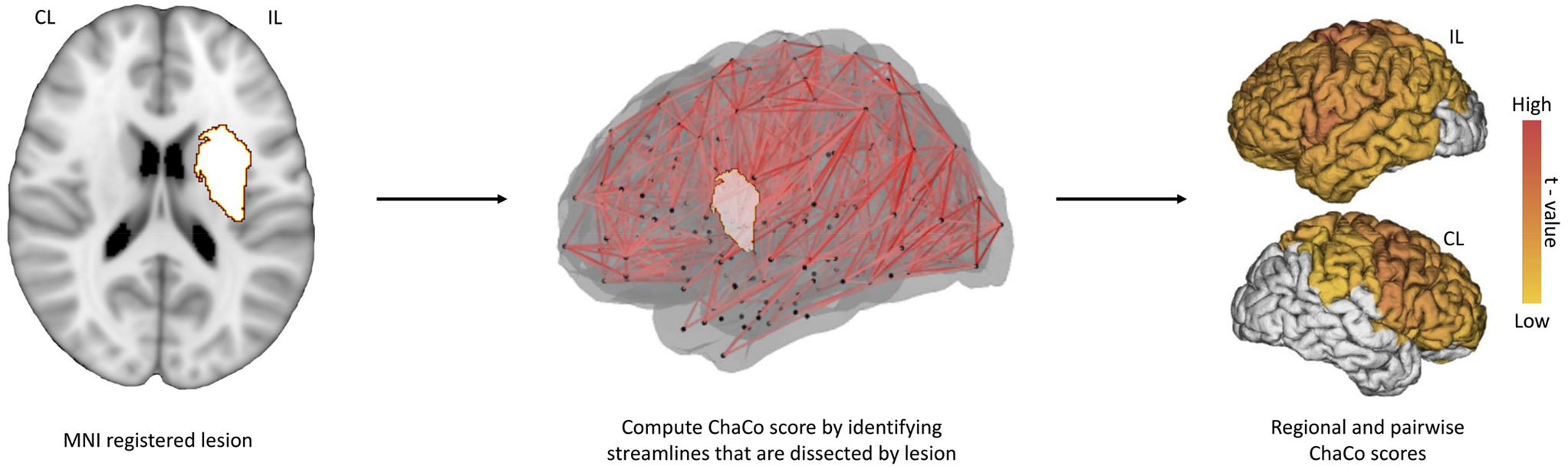
Quantification of structural disconnection: Binary lesions masks in MNI space are utilized to compute regional and pairwise region-to-region change of connectivity (ChaCo) scores by overlaying the lesion on a normative structural connectome. The ChaCo scores are computed by calculating the number of intersected streamlines on a regional (total % of disconnection for a given brain area) and pairwise (total % disconnection between a pair of brain regions) level. ChaCo scores were computed for 246 regions in the Brainnetome atlas. Regions exhibiting skewed data distribution (either close to 0 or 100% ChaCo) were not considered in the final analysis. Structural disconnection of 100 regions primarily belonging to the ipsilesional hemisphere were statistically analyzed (see Suppl. Figure 1). CL contralesional, IL ipsilesional, MNI Montreal Institute of Neurology, ChaCo change of connectivity.

### Statistical analysis

Statistical analyses were carried out in R version 4.2.3 ^33^. In the first step, 100 ordinal logistic regression models (function *polr* from the MASS package) were fitted to relate the regional structural disconnection value obtained at T_1_ to mRS at T_2_^34^. Models were adjusted for lesion volume, NIHSS at T_1_ and age. Lesion volumes were LOG_10_-transformed to improve data distribution. To address multicollinearity, we included lesion volume and NIHSS at T_1_ after residualization against the regional disconnection, in line with recent studies ^27,28^. Models were corrected for multiple comparisons via the Bonferroni-Holm false-discovery-rate (FDR) method and a leave-one-out analysis (LOOA) was applied to test the robustness of significant regional findings. Adjusted odds ratios (OR) for a one-unit increase of 0.01 in the ChaCo value are given for significant models with respective 95% confidence intervals and P_FDR_ values. OR values >1 indicate a higher risk of scoring 1 point higher on mRS at T_2_. In those regions whose overall level of disconnection was linked to the outcome after stroke, a pairwise region-to-region disconnectome analysis was added to explore which connections or networks might drive the statistical contribution of that region in the first step outcome model. Again, connections exhibiting skewed data were omitted from this second analysis step. Hence, for each region, up to 245 models (connections between the region and the remaining 245 regions of the Brainnetome atlas) were fitted. FDR-correction for multiple testing and LOOA were applied. Statistical significance was set at P_FDR_<0.05.

### Data availability

Data will be made available by authors upon reasonable request to the corresponding author.

## Results

### Demographics and clinical data

Table 1 shows the individual demographic and clinical data. The cohort comprised 38 severely affected stroke patients with supratentorial lesions [19 females, 23 right-hemispheric strokes, 2 left-handed, median age 73 years]. The median NIHSS score was 9.5 at T_1_. Median mRS was 4 at T_1_ and 3 at T_2_. Mean lesion volume was 58.25mL with a standard deviation of 70.01. A heatmap visualizing the distribution of the MNI-registered binary stroke lesions is shown in Figure 2.

**Table 1:** Demographic overview. Asterisks * in mRS T_2_ indicates that follow-up data was available after six months.

| ID | Study | Age range | Sex | Lesion side | Lesion Volume, mL | NIHSS T<sub>1</sub> | mRS T<sub>2</sub> | Thrombolysis | Thrombectomy | TICI |
| --- | --- | --- | --- | --- | --- | --- | --- | --- | --- | --- |
| 1 | C1 | 46-50 | Female | Left | 53.80 | 10 | 2 | Yes | Yes | NA |
| 2 | C1 | 71-75 | Female | Left | 5.82 | 9 | 4 | No | No | No |
| 3 | C1 | 66-70 | Male | Left | 6.57 | 8 | 3 | No | No | No |
| 4 | C1 | 81-85 | Male | Left | 1.73 | 4 | 3 | No | No | No |
| 5 | C1 | 46-50 | Male | Left | 24.44 | 7 | 2 | Yes | Yes | 3 |
| 6 | C1 | 86-90 | Female | Left | 0.99 | 1 | 1 | No | No | No |
| 7 | C1 | 41-45 | Male | Right | 79.82 | 13 | 2 | Yes | Yes | 3 |
| 8 | C1 | 56-60 | Male | Right | 2.53 | 13 | 4 | No | No | No |
| 9 | C1 | 66-70 | Male | Right | 25.09 | 3 | 1 | Yes | Yes | 3 |
| 10 | C1 | 71-75 | Female | Right | 26.76 | 3 | 1 | Yes | Yes | NA |
| 11 | C1 | 56-50 | Male | Right | 25.52 | 7 | 2 | Yes | Yes | 2B |
| 12 | C1 | 76-80 | Female | Right | 9.1 | 8 | 3 | No | No | No |
| 13 | C1 | 66-70 | Female | Right | 74.36 | 5 | 1 | Yes | Yes | 2A |
| 14 | C1 | 81-85 | Female | Right | 16.75 | 7 | 4 | No | No | No |
| 15 | C1 | 46-50 | Male | Right | 2.58 | 6 | 3 | Yes | No | No |
| 16 | C1 | 46-50 | Male | Right | 50.06 | 4 | 1 | Yes | Yes | 3 |
| 17 | C2 | 78-80 | Male | Left | 58.12 | 17 | 5* | No | No | No |
| 18 | C2 | 56-60 | Female | Left | 45.24 | 18 | 4 | NA | NA | NA |
| 19 | C2 | 81-85 | Female | Left | 101.41 | 20 | 6 | No | No | No |
| 20 | C2 | 81-85 | Male | Left | 3.60 | 8 | 3* | NA | NA | NA |
| 21 | C2 | 61-65 | Male | Left | 55.82 | 13 | 1 | No | No | No |
| 22 | C2 | 71-75 | Female | Left | 14.40 | 9 | 3 | Yes | Yes | 2B |
| 23 | C2 | 76-80 | Female | Left | 20.52 | 11 | 4* | No | Yes | 2B |
| 24 | C2 | 76-80 | Female | Left | 33.61 | 10 | 3 | No | No | No |
| 25 | C2 | 71-75 | Male | Left | 303.32 | 24 | 5* | Yes | Yes | 2B |
| 26 | C2 | 66-70 | Female | Right | 5.02 | 9 | 1 | NA | NA | NA |
| 27 | C2 | 76-80 | Male | Right | 100.96 | 11 | 3 | Yes | Yes | 3 |
| 28 | C2 | 71-75 | Male | Right | 75.22 | 15 | 6 | No | Yes | NA |
| 29 | C2 | 71-75 | Female | Right | 286.75 | 11 | 4* | Yes | No | No |
| 30 | C2 | 71-75 | Female | Right | 38.38 | 9 | 3* | Yes | Yes | 2A |
| 31 | C2 | 56-60 | Male | Right | 97.96 | 13 | 5 | Yes | Yes | NA |
| 32 | C2 | 66-70 | Female | Right | 7.36 | 11 | 1* | Yes | No | No |
| 33 | C2 | 76-80 | Male | Right | 108.41 | 16 | 6 | Yes | Yes | 0 |
| 34 | C2 | 76-80 | Female | Right | 120.41 | 8 | 4* | Yes | Yes | 2B |
| 35 | C2 | 81-85 | Female | Right | 33.49 | 15 | 5 | No | Yes | 2B |
| 36 | C2 | 76-80 | Male | Right | 178.07 | 17 | 4 | Yes | Yes | 3 |
| 37 | C2 | 71-75 | Female | Right | 27.56 | 5 | 1 | Yes | Yes | 2A |
| 38 | C2 | 76-80 | Male | Right | 91.82 | 15 | 4 | Yes | Yes | NA |

**Figure 2:**
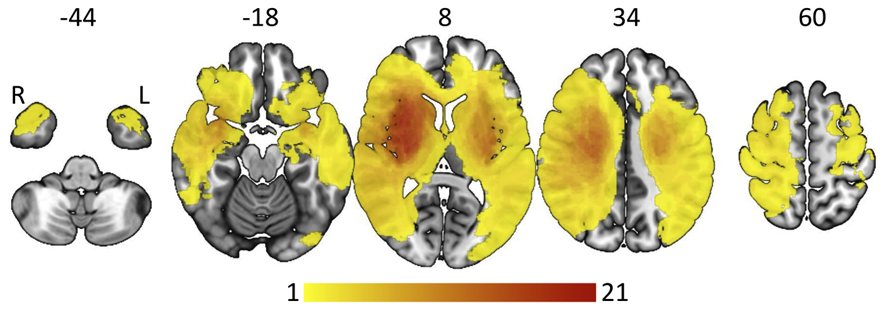
Lesion heatmap. The number of patients with lesioned voxel is color-coded with red colors indicating high overlap between lesions. Numbers above the maps indicate the z-value (slice) in MNI space. R right, L left.

### Regional disconnection and outcome after stroke

Ordinal logistic regression modelling revealed positive associations between lesion-induced regional disconnections primarily affecting frontal, parietal and temporal brain areas, and worse outcome after stroke (Figure 3A, Table 2). Highest OR values were computed for the precentral and postcentral gyrus. As an example, stacked histograms of mRS at T_2_ are plotted for the level of disconnection of the primary motor cortex (after median split of the patients into groups exhibiting lower and higher ChaCo values for illustration purposes, Figure 3B). Any of the 17 subcortical regions tested showed significant associations.

**Table 2:**
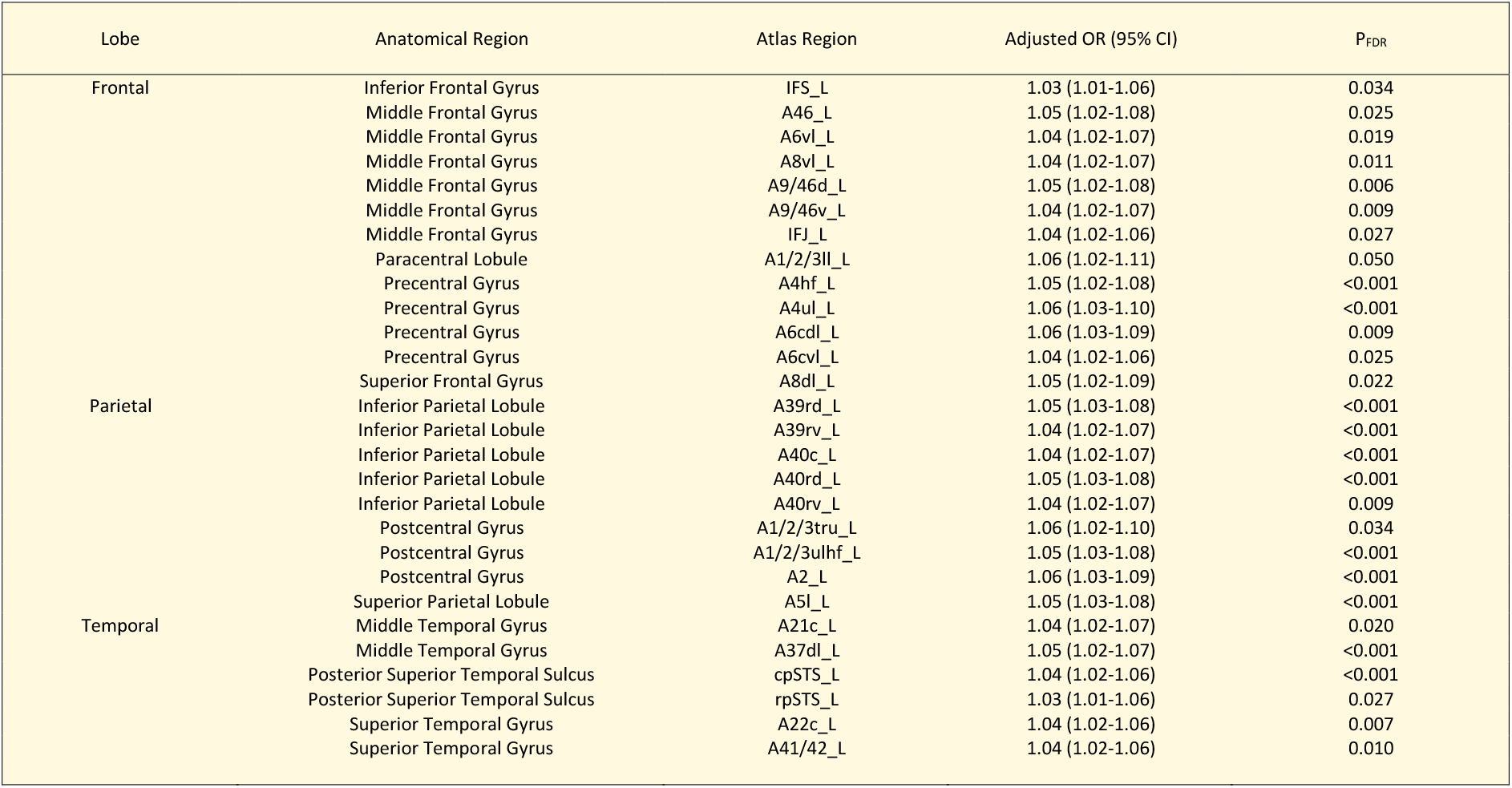
Results of ordinal logistic regressions between regional structural disconnection of 28 cortical brain regions and outcome after severe stroke. Results are corrected for age, lesion volume and NIHSS at T_1_. Adjusted OR with CI 95% and P_FDR_ values are given. OR was adjusted so that a one unit increase in the ChaCo value would relate to an increase of 0.01. _L left hemisphere, corresponding to the ipsilesional hemisphere. OR odds ratio.

**Figure 3:**
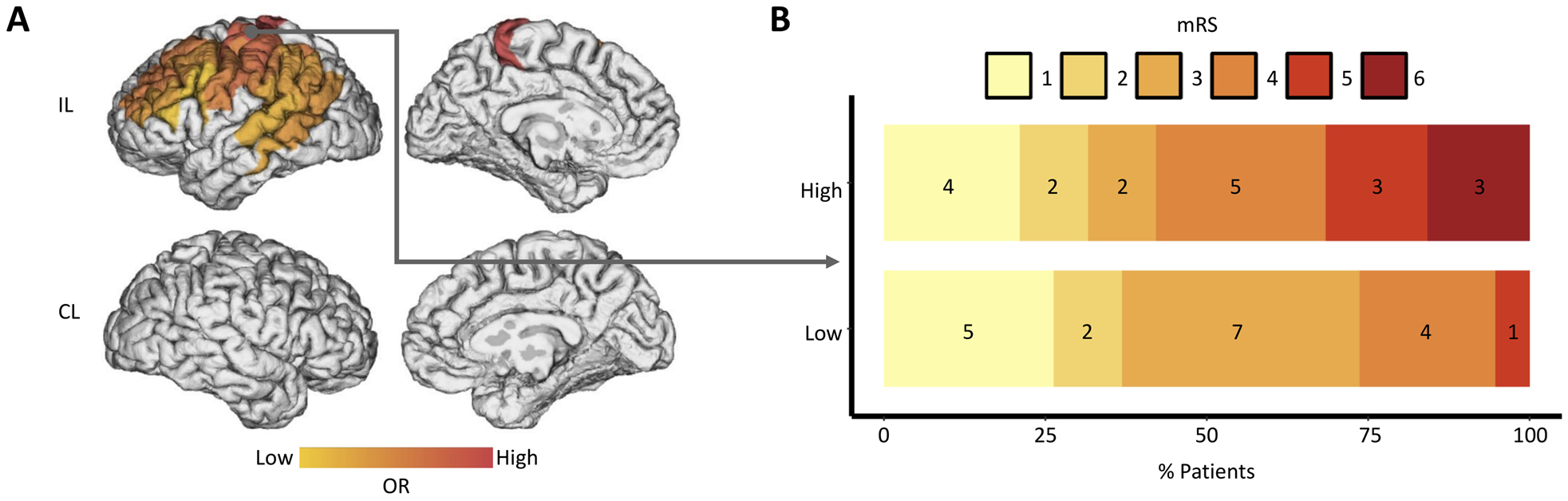
**A** Surface visualization of ORs corresponding to the significant associations between regional disconnection and outcome. Results are corrected for age, lesion volume and NIHSS at T_1_. Non-colorized regions were either excluded due to a skewed data distribution or lacked a significant association. IL ipsilesional, CL contralesional, OR odds ratio. **B** Shift plot illustrating the dichotomized structural disconnection of one representative Brainnetome region “A4ul_L” which represents the upper limb area of the left primary motor cortex. Outcome distribution is displayed for high and low disconnection values in yellow and red colors indicating lower and higher mRS scores, respectively.

### Pairwise disconnection patterns and outcome after stroke

Pairwise disconnection analyses provided additional insights into possible drivers of significant disconnectome-outcome relationships on a pathways or region-to-region level, results are depicted in Figure 4. The primary finding was the involvement of sparse intrahemispheric disconnections in frontal and prefrontal areas. Only labels contributing to the primary motor and dorsolateral premotor cortices showed an involvement of stronger frontoparietal and interhemispheric disconnectivity. Similarly, also temporal regional disconnections were driven by rather sparse intrahemispheric pathways except posterior middle temporal areas. In contrast to these frontal and temporal brain regions, the importance of parietal cortical disconnectivity for the outcome appeared to be driven not only by the structural state of extensive strong frontoparietal intrahemispheric, but also extensive interhemispheric connections. On visual inspection, these connections appeared to be most pronounced for posterior parietal cortices and inferior temporal cortices (Figure 4).

**Figure 4:**
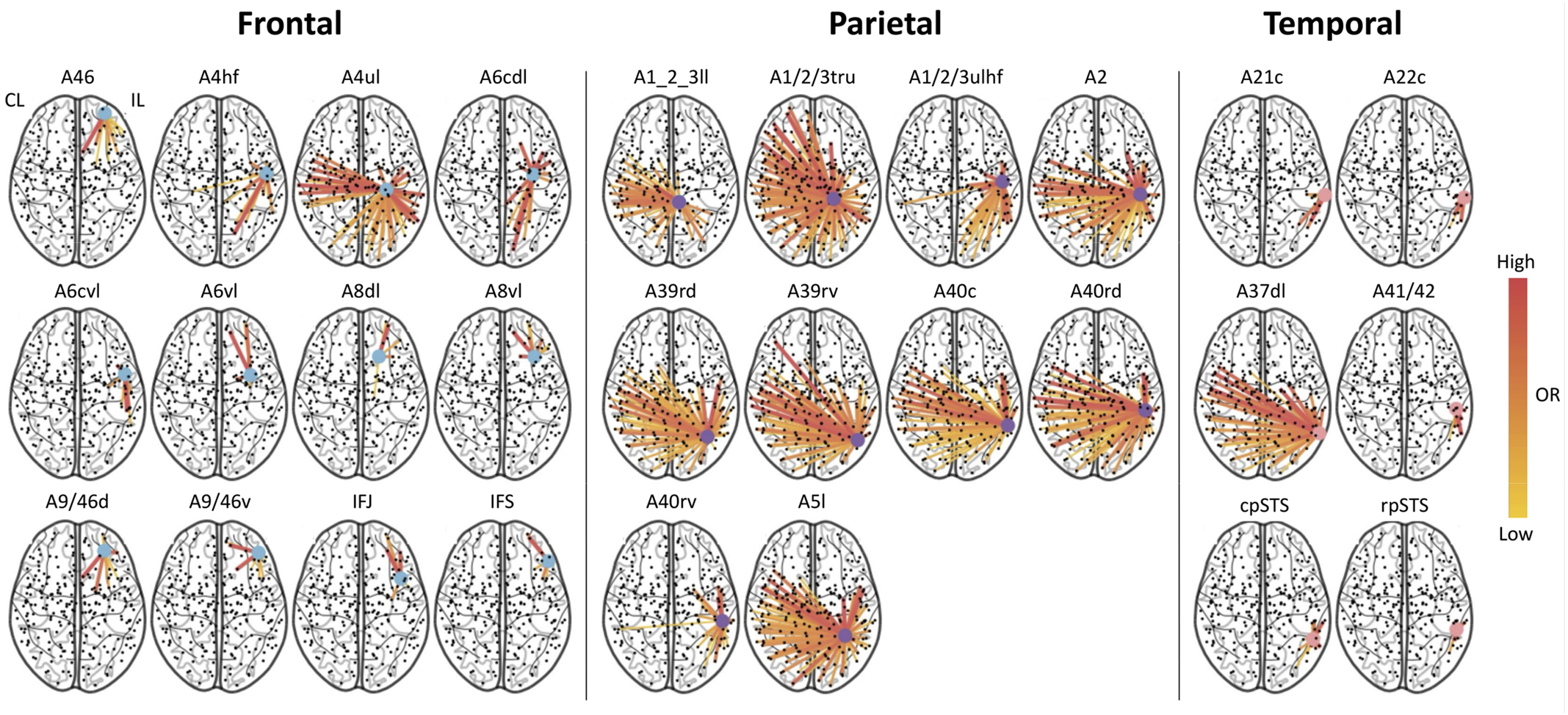
Glassbrain plots showing significant pairwise region-to-region disconnection profiles for every significant regional disconnection per lobe. Edges which showed a significant structure-outcome relation are colorized according to their adjusted OR values. Frontal and temporal regions exhibit a predominantly intrahemispheric disconnection profile while parietal regions show a strong edge disconnection not only to frontal areas of the ipsilesional but also to frontal and parietal brain areas of contralesional hemisphere. Results are corrected for age, lesion volume and NIHSS at T_1_. Adjusted OR are color-coded. List of region names can be found in the Suppl. Table 1. CL contralesional, IL ipsilesional, OR odds ratio.

## Discussion

The main finding of the present study was that lesion-induced disconnections of frontal, parietal and temporal cortical brain areas, adjusted for the initial deficit, lesion volume, and age, are significantly associated with outcome after severe stroke. The spatial distribution of the underlying pathways mediating the disconnection-outcome association was not diffuse or unspecific. It revealed location specific results: For frontal, prefrontal and most temporal brain areas, it was primarily driven by relatively sparse intrahemispheric disconnections. In contrast, the ipsilesional primary motor cortex, the dorsal premotor cortex, and various parietal brain regions showed a remarkable involvement of either frontoparietal intrahemispheric or additionally interhemispheric disconnections. These results indicate that localized disconnection of multiple regions embedded in the structural frontoparietal network correlates with outcome after severe stroke, and that primary motor and parietal cortices might gain a particular importance as they structurally link frontoparietal networks of both hemispheres.

These findings extend previous data regarding the functional importance of lesion-induced disconnectome profiles for outcome inference after stroke. One study mapped cognitive functions, basic mobility, and daily activity, assessed after six months, to disconnectomes in 40 acute stroke patients. In that cohort with rather moderate symptom burden, functional scores were primarily predicted by intrahemispheric network disconnections. These were localized in the frontal lobes for applied cognitive tests and basic mobility, and in frontal and frontoparietal networks for daily activities, respectively. Notably, for the latter, a pairwise region-to-region approach was found to be particularly informative for outcome prediction when compared to lesion volumes or regional disconnectivity values alone ^12^. In contrast to that study, the present cohort included more severely impaired patients. The current analyses suggest that interhemispheric parietal disconnectivity might play a crucial role in the recovery of stroke patients with severe impairments, though variations exist in methodologies, such as statistical approaches or clinical assessment tools utilized.

A recent study that derived connectome data from tract-related microstructure and aimed at predicting the natural recovery in acute stroke patients corroborates our findings ^14^. The authors reported that connections within the parietal cortex including the intraparietal sulcus as well as the superior parietal gyrus on both hemispheres, with a specificity of the unaffected hemisphere for severely impaired patients, improved prediction of recovery ^14^. Various electrophysiological, functional, and structural imaging studies have already referred that interhemispheric connections and the contralesional hemisphere might be particularly important for stroke recovery of severely impaired patients. For instance, stimulation studies have revealed that the suppression of regional excitability ^35^ and the perturbation of information processing ^36,37^ on the contralesional hemisphere could impair motor functions in chronic stroke patients. Functional imaging has indicated widespread increases in contralesional brain activation from frontal to occipito-parietal brain areas ^22,38,39^, particularly after severe stroke. It has been argued that attentional processes or adaptive motor learning strategies to compensate for motor impairment might drive such changes in activation ^40-42^. Coupling analyses based on resting-state functional data found that interhemispheric rather than intrahemispheric connection profiles were significantly related to acute deficits ^43^ and later outcome ^44,45^. Another study investigated 18 chronic stroke patients and reported that patients with a good outcome exhibited a stronger interhemispheric parietal coupling between the anterior intraparietal sulcus than patients with a poor outcome ^46^. Structural analyses have complemented this largely coherent picture. For instance, one study showed that contralesional cortical thickness assessed early after severe stroke could inform about subsequent recovery^27^. Another study found that white matter disconnections affecting parietal cortices and the posterior corpus callosum were correlated with motor rehabilitation outcome in chronic stroke patients ^16^.

Apart from interhemispheric connections, the importance of the regional disconnection of the primary motor cortex, the dorsal premotor cortex, and multiple parietal cortices along the intraparietal sulcus was additionally driven by intrahemispheric reciprocal frontoparietal disconnectivity. This finding corroborates studies which already emphasized that ipsilesional frontoparietal network integrity, characterized by regional structural or connectivity analyses, is an important factor for a favourable outcome. Of note, this might particularly apply to severe stroke ^22^ as severity-specific roles of frontoparietal network involvement were statistically indicated by subgroup analyses ^47^ or, at least with a strong statistical trend, by previous interaction modelling ^48^.

Collectively, when considering both inter- and intrahemispheric findings, particularly in the parietal regions, the current disconnectome data emphasize that parietal brain regions may hold particular significance for recovery after severe stroke. This is because they are involved, at least structurally, in extensive frontoparietal networks that span both hemispheres.

Interestingly, only sparse interhemispheric disconnections mediated the behavioural effect of most frontal brain regions, except for the upper limb area of the ipsilesional primary motor cortex. At least from this structural disconnectome perspective, secondary motor areas of the frontal lobe might therefore be primarily involved in recovery processes after stroke via intrahemispheric connections. This interpretation would find compelling parallels in various functional connectivity studies reporting changes in intrahemispheric premotor-motor couplings and significant coupling-outcome relationships more frequently than interhemispheric connections. Consistent and functionally relevant alterations in interhemispheric connectivity were found for the primary motor cortices of both hemispheres. Preserved or increased connectivity was positively related to a better outcome, in the general stroke population ^49^, and patients with severe hand weakness ^50^. In agreement, previous structural studies ^4^ have linked interhemispheric connectivity with cortico-spinal tract damage, bilateral brain activation and motor impairment ^51,52^ or functional improvement during motor rehabilitation ^53^. The present structural disconnectome study adds cross-sectional imaging and longitudinal clinical data to this existing research and underlines that the primary motor cortex does not act solely as a key area within the ipsilesional motor network, but also as a crucial hub maintaining functionally important structural connections to the contralesional hemisphere. Their disturbances impede recovery after severe stroke beyond otherwise important influential factors such as initial symptom burden and mere lesion volume.

Given these localized disconnectome-outcome relationships in severe stroke, the question emerges how such insights can add to stroke recovery research. One idea arises from recent ambitions to develop non-invasive brain stimulation towards patient-tailored treatment strategies ^24,54^ to overcome difficulties in translating *one-suits-all* approaches from neuroscience to clinical applications ^55,56^. For instance, patients with preserved interhemispheric connections between primary motor cortices or parietal brain regions might be particularly susceptible for bilateral brain stimulation. First attempts to uncover possible patterns suitable for patient stratification have been recently published in very small cohorts for bilateral motor cortex stimulation combined with motor therapy ^53^ and contralesional parietal cortex stimulation ^46^ in chronic stroke.

There are several important limitations to note. First, the cohort comprised severely impaired stroke patients with rather large lesions primarily localizing in the vascular territory of the middle cerebral artery. This introduces a systematic bias which is likely to explain (1) a relevant portion of the disconnectome profiles, both on the regional and region-to-region level and (2) why any subcortical region, of which all showed very high disconnection values, could be linked to outcome variability due to reduced variance across the individual patients. Second, all stroke lesions were flipped to the left hemisphere. Disconnectome values in the contralesional hemisphere exhibited either a too high skew or low skew and were thus not eligible for statistical analysis. Only 100 regions were included in the first step of analysis. A larger sample size might result in a better data distribution on both hemispheres which might in turn lead to the detection of further disconnection-outcome associations, particularly in the light of potentially more sensitive hemisphere specific analyses^57^. Third, the present cohort comprised a relatively small group of early subacute ischemic stroke patients. Regression models were corrected for multiple testing, and a LOOA was additionally carried out to improve specificity of the results at the cost of reduced sensitivity. The analyses were correlative in nature. Future studies combining larger samples and predictive modelling including out-of-sample validation would be needed to further test the robustness of the present findings. Finally, follow-up clinical data were acquired in the late subacute stage of recovery. The primary outcome was global disability, operationalized by mRS, dominated by motor functions. Hence, to what extent the findings might generalize to other time points or to other more precise functional clinical scores or domains such as language or cognition remain interesting topics for upcoming studies.

## Sources of Funding

This work was funded by the Deutsche Forschungsgemeinschaft (DFG, German Research Foundation) SFB 936 – 178316478, projects C1 to C.G., C2 to G.T. and SFB TRR169 project A3 with the National Science Foundation of China (NSFC) in project Crossmodal Learning to C.G., and the Else Kröner-Fresenius-Stiftung (2016_A214 to R.S.). R.S. and C.U.C. are supported by an Else Kröner Exzellenzstipendium from the Else Kröner-Fresenius-Stiftung (2020_EKES.16 to R.S., 2018_EKES.04 to C.U.C.). F.Q. is supported by the Gemeinnützige Hertie-Stiftung (Hertie Network of Excellence in Clinical Neuroscience). Open Access funding enabled and organized by Projekt DEAL.

## Statement of ethical approval

The original studies were conducted in line with the ethical declaration of Helsinki and were granted permission by the local ethics committee of the Chamber of Physicians Hamburg. All participants or their legal guardian provided informed consent.

## Notes

### Competing Interest Statement

The authors have declared no competing interest.

### Author Declarations

The original studies which were the base for this secondary analysis were conducted in line with the ethical declaration of Helsinki and were granted permission by the ethics committee of the Chamber of Physicians Hamburg. All participants or their legal guardian provided informed consent.

