## Supplementary Material for "Frontoparietal structural network disconnections correlate with outcome after a severe stroke"

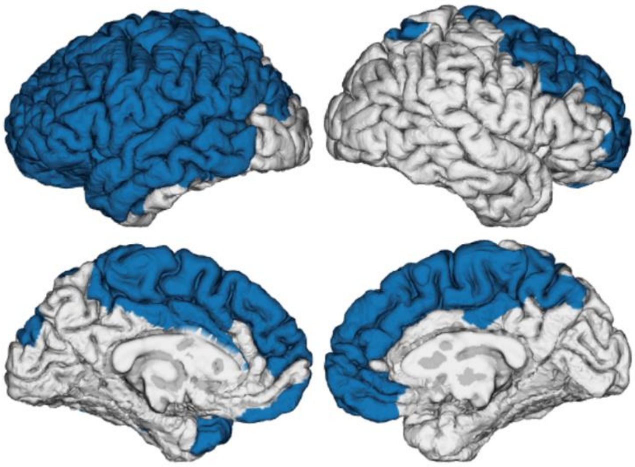

**Supplementary Figure 1:** Colorized in blue are all cortical brain areas that were included into further analysis after skewed data removal at a threshold of skew either >1.3 or <-1.3.

**Supplementary Table 1:** Listed are all brain areas that were included into further analysis after skewed data removal.

| Region | Anatomical name |
|---|---|
| LAMYG_L | Amygdala |
| MAMYG_L | Amygdala |
| DCA_L | Basal Ganglia |
| DLPU_L | Basal Ganglia |
| GP_L | Basal Ganglia |
| NAC_L | Basal Ganglia |
| VCA_L | Basal Ganglia |
| VMPU_L | Basal Ganglia |
| A23C_L | Cingulate Gyrus |
| A23C_R | Cingulate Gyrus |
| A24RV_L | Cingulate Gyrus |
| RHIPP_L | Hippocampus |
| A44D_L | Inferior Frontal Gyrus |
| A44OP_L | Inferior Frontal Gyrus |
| A44V_L | Inferior Frontal Gyrus |
| A45C_L | Inferior Frontal Gyrus |
| A45R_L | Inferior Frontal Gyrus |
| IFS_L | Inferior Frontal Gyrus |
| A39RD_L | Inferior Parietal Lobule |
| A39RV_L | Inferior Parietal Lobule |
| A40C_L | Inferior Parietal Lobule |
| A40RD_L | Inferior Parietal Lobule |
| A40RV_L | Inferior Parietal Lobule |
| DIA_L | Insular Gyrus |
| DID_L | Insular Gyrus |
| G_L | Insular Gyrus |
| VIA_L | Insular Gyrus |
| VID/VIG_L | Insular Gyrus |
| A10L_L | Middle Frontal Gyrus |
| A10L_R | Middle Frontal Gyrus |
| A46_L | Middle Frontal Gyrus |
| A46_R | Middle Frontal Gyrus |
| A6VL_L | Middle Frontal Gyrus |
| A6VL_R | Middle Frontal Gyrus |
| A8VL_L | Middle Frontal Gyrus |
| A8VL_R | Middle Frontal Gyrus |
| A9/46D_L | Middle Frontal Gyrus |
| A9/46D_R | Middle Frontal Gyrus |
| A9/46V_L | Middle Frontal Gyrus |
| IFJ_L | Middle Frontal Gyrus |
| A21C_L | Middle Temporal Gyrus |
| A21R_L | Middle Temporal Gyrus |
| A37DL_L | Middle Temporal Gyrus |
| ASTS_L | Middle Temporal Gyrus |
| A11L_L | Orbital Gyrus |
| A11M_L | Orbital Gyrus |
| A12/47L_L | Orbital Gyrus |

| | |
|---|---|
| A12/47O_L | Orbital Gyrus |
| A1/2/3LL_L | Paracentral Lobule |
| A1/2/3LL_R | Paracentral Lobule |
| A4LL_L | Paracentral Lobule |
| A4LL_R | Paracentral Lobule |
| A1/2/3TONIA_L | Postcentral Gyrus |
| A1/2/3TRU_L | Postcentral Gyrus |
| A1/2/3ULHF_L | Postcentral Gyrus |
| A2_L | Postcentral Gyrus |
| CPSTS_L | Posterior Superior Temporal Sulcus |
| RPSTS_L | Posterior Superior Temporal Sulcus |
| A4HF_L | Precentral Gyrus |
| A4T_L | Precentral Gyrus |
| A4T_R | Precentral Gyrus |
| A4TL_L | Precentral Gyrus |
| A4UL_L | Precentral Gyrus |
| A6CDL_L | Precentral Gyrus |
| A6CDL_R | Precentral Gyrus |
| A6CVL_L | Precentral Gyrus |
| A5M_L | Precuneus |
| A5M_R | Precuneus |
| A7M_R | Precuneus |
| A10M_R | Superior Frontal Gyrus |
| A6DL_L | Superior Frontal Gyrus |
| A6DL_R | Superior Frontal Gyrus |
| A6M_L | Superior Frontal Gyrus |
| A6M_R | Superior Frontal Gyrus |
| A8DL_L | Superior Frontal Gyrus |
| A8DL_R | Superior Frontal Gyrus |
| A8M_L | Superior Frontal Gyrus |
| A8M_R | Superior Frontal Gyrus |
| A9L_L | Superior Frontal Gyrus |
| A9L_R | Superior Frontal Gyrus |
| A9M_L | Superior Frontal Gyrus |
| A9M_R | Superior Frontal Gyrus |
| A5L_L | Superior Parietal Lobule |
| A7PC_L | Superior Parietal Lobule |
| A7R_L | Superior Parietal Lobule |
| A7R_R | Superior Parietal Lobule |
| A22C_L | Superior Temporal Gyrus |
| A22R_L | Superior Temporal Gyrus |
| A38L_L | Superior Temporal Gyrus |
| A38M_L | Superior Temporal Gyrus |
| A41/42_L | Superior Temporal Gyrus |
| TE1.0/TE1.2_L | Superior Temporal Gyrus |
| CTTHA_L | Thalamus |
| LPFTHA_L | Thalamus |
| MPFTHA_L | Thalamus |
| MPMTHA_L | Thalamus |
| OTHA_L | Thalamus |
| PPTHA_L | Thalamus |
| RTTHA_L | Thalamus |
| STHA_L | Thalamus |

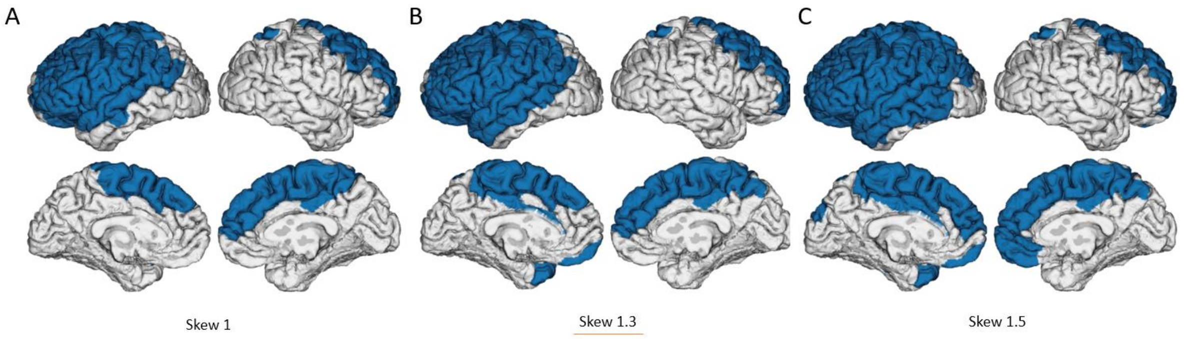


**Supplementary Figure 2:** Influence of skewness threshold. Alternative thresholds at 1.0 (A) and 1.5 (B) leading to other subsets of regions for further statistical analyses are presented. B indicates the threshold at 1.3 which served for the primary analysis (Depicted in Supp. Fig. 1). Please note that alterative thresholds would largely lead to comparable sets of regions.
